# Smartphone Apps for Cannabis Cessation: Quality Assessment and Content Analysis

**DOI:** 10.1101/2023.09.27.23296238

**Authors:** Siddharth Seth, Sumedha Kushwaha, Reshma Prashad, Michael Chaiton

**Affiliations:** Queen’s University, Kingston, Canada; Centre for Addiction and Mental Health, Toronto, Canada; Institute of Medical Science, University of Toronto, Canada; Longo Faculty of Business, Humber College, Canada; Dalla Lana School of Public Health, University of Toronto, Canada

**Keywords:** cannabis, cessation, mHealth interventions, mobile applications

## Abstract

**Background:** The rising rates of cannabis use, cannabis-related problems, and hospitalization rates related to cannabis usage warrant further research into better treatment methods and interventions.

**Purpose:** This study aims to identify the quality of free cannabis cessation applications available on both the Apple App Store and the Google Play Store and analyse their features, content, and adherence to evidence-based practices.

**Methods:** A systematic search was conducted in April 2023 using a variety of keywords. The applications were deemed eligible if they were free, in English, available on both the Apple App Store and the Google Play Store and were related to cannabis cessation. Each application was used for at least one month and were rated on the Mobile App Rating Scale by two users. There was excellent agreement determined between the two reviewers (≤2 points on all categories).

**Results:** A total of 4 applications were included in the overall quality and content analysis. The mean total quality scores of applications were determined to be 3.36 out of 5 indicating a poor to acceptable quality of applications. It was determined that there was a very limited number of available applications for users and those that were available were not of high quality with few applications incorporating evidence-based practices.

**Conclusions:** There are a very limited number of cannabis cessation applications available, and the currently available products have poor to acceptable quality.

## INTRODUCTION

On October 17, 2018, Canada legalized recreational cannabis through the Cannabis Act, making it the second country in the world, after Uruguay, to do so at the national level.^1^ Initially passed to protect youth health, enhance public awareness of cannabis-related health risks, and provide access to a quality-controlled supply of cannabis, the legislation appears to have led to a marked increase in cannabis usage across Canada.^2,3^ The 2021 Canadian Cannabis Survey indicated that 17% of Canadians aged 16 and older reported using cannabis in the past 30 days, with an average consumption of 14.3 days.^3^ Another study on cannabis legalization from 2001 to 2019 revealed significant increases in daily cannabis use, past 12-month use, and cannabis-related problems.^4^

A study combining 19 iterations of the Centre for Addiction and Mental Health (CAMH) Monitor Surveys using a pre-post design found that youth and young adults had the highest rates of cannabis-related problems, such as dependence, anxiety and depression disorders, and physical health effects. Although younger individuals continue to exhibit higher levels of cannabis use, the largest rise in consumption since legalization was observed among middle-aged and older adults (those aged 45-64 and 65 and above).^5^ The study concluded that the Cannabis Act was strongly associated with increased likelihood of cannabis use, daily cannabis use, and cannabis-related problems, including dependence. This finding is supported by Ontario Cannabis Store reports stating that total cannabis sales in Ontario increased by 182% (from 35 million grams to 99 million grams) between the first and second fiscal years.^5.6^

According to the Canadian Institute for Health Information, cannabis was the leading contributor to hospitalization rates for substance use among youth aged 10-24 (40%) between 2017-2018. For those aged 25 and older, cannabis was associated with 11% of hospitalizations. Given the age-standardized rate of 104 individuals per 100,000 population, it is clear that significant resources are required.^7^ Nonetheless, addiction treatment wait times have increased over the last decade, with the average wait time for residential programs reaching 50 days by 2019. Many clients were reported to have been hospitalized, jailed, attempted suicide, or died while awaiting treatment.^8^

Various methods can be employed to facilitate cannabis addiction treatment, such as cognitive-behavioural therapy, which aids individuals in identifying and correcting problematic behaviours, including addiction. Contingency management is another approach that monitors target behaviours and provides or removes tangible rewards to reinforce positive behaviours. Although no medications are currently approved for cannabis therapy, some are being trialled to help control withdrawal symptoms, and newer clinical trials are focusing on allosteric modulators to inhibit cannabis’s rewarding effect.^9^

Mobile phones have revolutionized communication technology, enabling users to access various applications for different purposes. With a global audience of 6.4 billion individuals as of 2021, this technology is pervasive and widely used.^10^ The literature documents the use of mobile applications for alcohol and tobacco therapy, and research suggests these applications may promote successful quit attempts among e-cigarette users or vapers.^11-13^ Available applications focus on habit tracking, therapy, or education. Given the role of this technology in disseminating cannabis-related news and information, the present study aimed to examine the quality of cannabis cessation apps, their content and features, popularity among users, and adherence to evidence-based practices. This study primarily focused on free English-language mobile applications available on the Apple App Store and Google Play Store.

## MATERIALS AND METHOD

### Search Strategy

Using the terms “cannabis cessation”, “cannabis quit”, “quit weed”, “marijuana stop”, “stop pot”, “weed cessation”, and “ganja”, the primary and secondary author searched for smartphone apps targeting Cannabis Cessation on the Canadian Apple and Google play stores in April 2023. Preliminary searches on the Apple App Store website and the embedded App Store app in iPhones showed drastically different sets of results. Based on the inclusion criteria, we selected four applications:

1. Grounded-Quit Weed
2. Marijuana Addiction Calendar
3. Marijuana Anonymous
4. Quit Weed

### Eligibility Criteria

Mobile applications related to cannabis cessation or cannabis behaviour modification were included. Applications that were not available for free download or not in English were excluded. Applications that were not available on both the Google Play Store and Apple App Store were also excluded.

Once the applications were removed through a preliminary analysis, the remainder of the applications were then examined by checking their description, profiles, whether they addressed cannabis cessation, and their functionality. Those that did not meet the required criteria were excluded. The remaining eligible applications (n=7) were then independently reviewed by the primary and secondary author through a 2-pronged screening approach. Firstly, the profiles of the apps were examined on the online app stores, and secondly, an ancestry search was conducted of developer websites, profiles, and overall online presence, if the information was available. Apps that were confirmed to have no association with cannabis cessation were excluded, resulting in a final sample of 4 applications.

### Assessment of Quality, Contents and Features, and Popularity Among Users

To ensure the accuracy and reliability of our results, each app was rigorously tested over a short period of time (up to 1 month). This allowed us to classify the applications based on quality, features, content, and perform the necessary evaluation of its tools. The popularity of the application was gauged through a variety of metrics including the number of downloads, the number of reviews, and the user ratings. These metrics provided a composite score by which the usage of the application could be estimated. We exclusively assessed the applications on Apple devices and did not evaluate them on Android devices.

The quality of each application was evaluated using the Mobile App Rating Scale (MARS), which is a multidimensional measure that is designed to rate the quality of mobile health applications.^14^ The MARS is comprised of five categories – engagement, functionality, aesthetics, information quality, and subjective quality – with a total of 23 collective items rated using a 5-point scale. For items that could not be adequately assessed, an option of “not applicable” was available. The MARS also contained a subjective quality category; however, it was not factored into the final score. To identify the strengths and weaknesses of each app, mean scores were calculated for each of the four objective categories, while the sum of all four scores provided an overall quality score for each app. However, noting the variability found among health-related apps, the Mobile App Rating Scale (MARS) does not provide a defined threshold for the type of scores a high-quality app should obtain on the scale. We based our analysis on a previous study that assessed the quality and content of mobile vaping cessation applications established a MARS total mean score of 3 as the cut-off for apps with acceptable quality.^13^

## Data Extraction and Quality Assessment

2 raters tested the MARS with a randomly selected app. Results from the test app showed substantial agreement with minor differences (≤2 points) that were discussed and resolved. The remaining applications were then independently evaluated and once again showed excellent agreement between the two reviewers (≤2 points).

## RESULTS

A total of 4 apps for cannabis cessation were identified and the app selection process is shown in Figure 1. Figure 2 provides a visual overview of the remaining 4 apps included in the analysis. Characteristics and quality scores of each app are presented in Table 1.

**Table 1:**
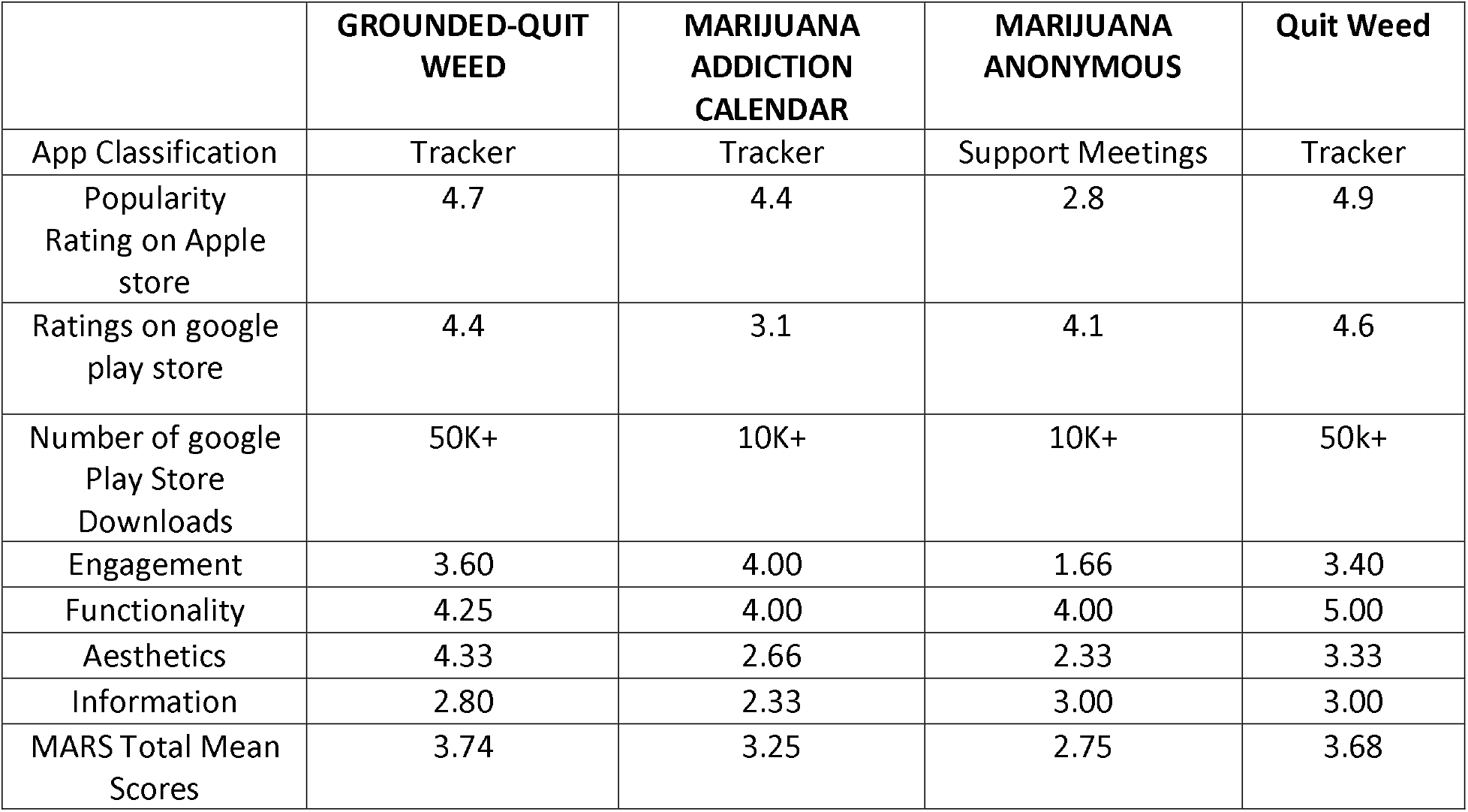
Characteristics and Quality Scores of the Applications.

**Figure 1.**
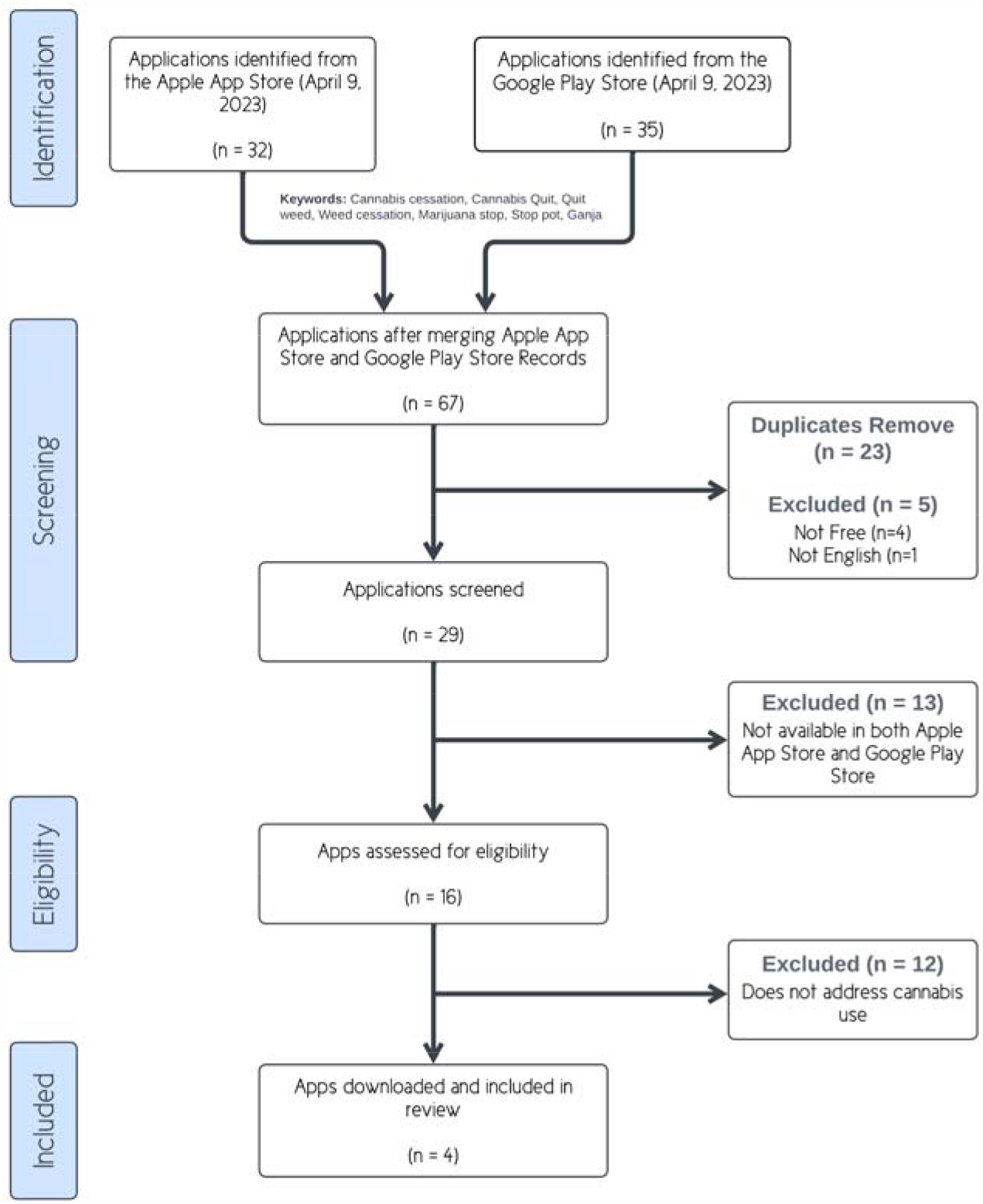
App Selection Process

**Figure 2.**
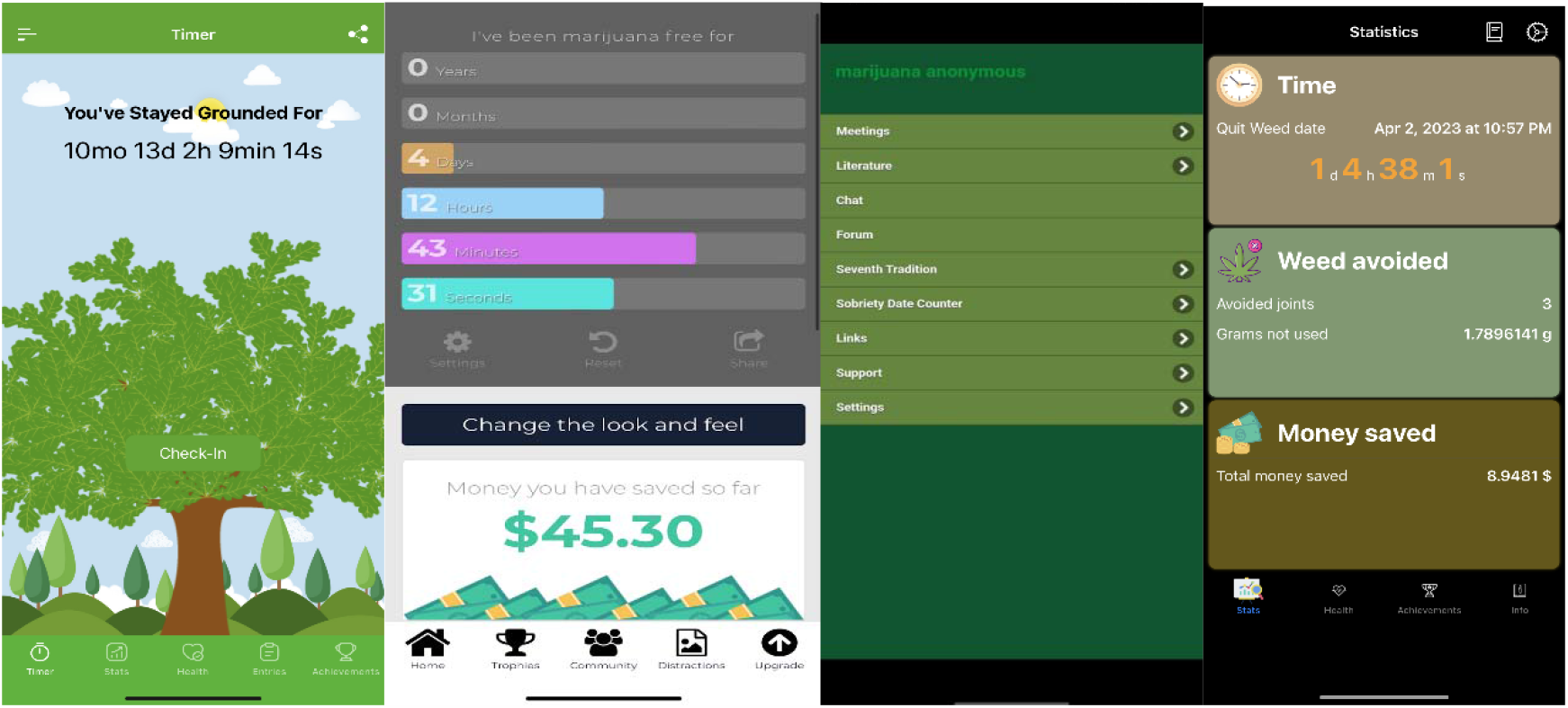
Visual Overview of the 4 mobile applications included in the analysis (from left to right: “Grounded – Quit Weed,” “Marijuana Addiction Calendar,” “Marijuana Anonymous,” and “Quit Weed”)

### Assessment of the Applications

The overall mean score for MARS app quality was 3.355, indicating poor to acceptable quality of the applications. Among the four applications, “Grounded - Quit Weed” received the highest MARS score of 3.74, while “Marijuana Anonymous” received the lowest score of 2.75.

On average, the functionality category received the highest mean score (4.3125), followed by engagement (3.165), aesthetics (3.1625), and information (2.7825) with the lowest score. These findings are summarized in Table 1.

Validity of the information inside the applications was done by conducting a literature review and cross-referencing this with the content of the application. This analysis noted that only some of the information provided by the applications was supported by strong scientific evidence. In accordance with the inclusion criteria, all four applications were available on both the Apple App Store and the Google Play Store.

### Features

The “Grounded - Quit Weed”, “Marijuana Addiction Calendar”, and the “Quit Weed” applications both utilized a similar approach, by using a timer to track the amount of time spent without cannabis use. The applications also provided some supporting data, to assist with tracking. These were classified as “tracking” applications. In contrast, the “Marijuana Anonymous” application did not offer any usage tracking but instead provided a networking platform to find licensed Marijuana Anonymous meetings and distribute spiritual writings and scriptures.

The tracking applications asks the user a series of questions regarding their cannabis use, specifically how many grams of cannabis they used per day, the price of each gram of cannabis, and the quit date. “Grounded – Quit Weed” and the “Marijuana Addiction Calendar” both asked questions about what the user’s goal was (quitting, tolerance break, or tracking usage). “Marijuana Anonymous” did not ask for any specific information but did include a category to document the amount of time spent sober. The usage tracking applications continue to track abstinence duration, money saved, and cannabis not smoked throughout the time period the application was used.

The “Grounded - Quit Weed” application provided users with a pseudo-diary where individuals were able to record their cravings, triggers, moods, how much money spent, and general notes. Nothing similar was seen in the other three applications. All three of the usage tracking applications contained a motivation/distraction centre. With “Grounded – Quit Weed”, this was called the “motivation” subsection and provided users with daily rotating quotes. The “Marijuana Addiction Calendar” application had a subsection titled “distractions.” This section included a limited selection of images, sounds, games, and workouts. “Quit Weed” also seemed to have a motivation section, but it was locked behind a pay wall. “Marijuana Anonymous” did not contain an equivalent.

The three applications with usage tracking features included a trophy/award/achievement section where individuals could receive badges for abstinence. Nothing similar was found in the “Marijuana Anonymous” application. The “Grounded – Quit Weed” application also contained a section where individuals could track their urges/intensity of urges in a table format to review. This data was collected from the pseudo-diary.

The “Grounded – Quit Weed” application provided limited information regarding the “phase” of the quitting cycle. It divided the quitting progress into three phases and provided information regarding the phase, common side effects an individual can experience, and the main focus an individual should have during the phase. This section also provided information on tips to stay off cannabis and a self-assessment in order to determine how much cannabis affected a user’s life. The “Quit Weed” application provided information about the adverse effects of cannabis usage and the duration it takes for users to overcome them after quitting. The duration was provided to users via a countdown. This information was well supported with citations and a source was provided for most side effects. Further information regarding recovery phases was hidden behind a paywall, thus its validity was unable to be evaluated. The “Marijuana Anonymous” application did not provide specific information regarding the quitting process but instead a range of documents that described the purpose of Marijuana Anonymous, the tenets of the organization, stories of individuals in the organization, and information about their 12-step process. It was noted that the documents and texts had heavy religious and biblical undertones. The “Marijuana Addiction Calendar” did not contain any similar content.

Two of the usage tracking applications both offered a community chat feature. The “Marijuana Addiction Calendar” allows individuals to post in the forum after signing in with Apple. The forum was not very active, with only a couple of users using this feature weekly. The discussions and posts typically centred on individuals’ initial attempts at quitting, documenting their setbacks, and seeking advice, strategies, and encouragement. Individuals are also able to like, reply to, or flag other people’s comments as inappropriate. However, this feature was limited in that with the free membership as individuals were limited in how much they can post. The application also has a secondary feature where individuals are also able to submit testimonials about the effectiveness of this application on their quitting journey. These testimonials are then displayed to all users. The “Grounded – Quit Weed” application also advertised a community chat feature; however, this was contained in its premium paid version and thus was not evaluated. The “Quit Weed” and “Marijuana Anonymous” applications contained no similar content.

It was important to note that these applications were aimed towards abstinence from cannabis use, either permanently or for a short period of time to build up a tolerance. Specifically, the description of the “Grounded - Quit Weed” application stated that it also assists individuals who want to take a “tolerance break” from cannabis.

### App Store Evaluation

All of the applications offered some sort of in-app purchases or had a donation feature. The “Grounded – Quit Weed” application seemed to offer the most amount of value for the price. The application offered a subscription service called “Grounded Plus” which ranged from $7.49 per month to $35.99 per year for access to a trigger tracker, a community centre, a withdrawal tracking system, biometric IDs, and motivational testimonies and quotes. Furthermore, the paid feature also provided access to entry insights, which allows users to analyse their feelings, coping mechanisms, and the people they’re with when they feel urges or when they smoke.

The “Marijuana Anonymous” application does not have any in-app purchases and instead asked for donations from members of the organization. The donations are expected to go to Marijuana Anonymous World Services.

The “Marijuana Addiction Calendar” application also offered a membership option. The price ranged from $6.49 a month to $52.99 for the entire year. This subscription provided users with the ability to add login information to the application, unlock all of the distractions, provide unlimited community posting, unlocking all custom skins, and removes all of the advertisements in the application.

The “Quit Weed” application offered a paid “PRO” version for $13.99 as a one-time payment that unlocked visualization of progress (with three recovery phases and further information regarding the quitting progress), a list of possible withdrawal symptoms and tips for each phase, as well as a motivation section. This paid version also removed all advertisements.

The pricing for all of the applications was only evaluated on the Apple App Store and in Canadian dollars. The paid features were not tested or trialled.

It was noted that the version of the applications on the Apple Store was more up to date and had been often more recently updated. However, the version number was not an accurate comparator between how up to date an application was. For example, the “Quit Weed” application had been newly added to the Apple App Store and thus had a lower version number, but the features were up to date than its Android equivalent. In the “Grounded - Quit Weed” and the “Marijuana Addiction Calendar” application, the more recently updated version was rated higher whereas the opposite was found for Marijuana Anonymous. The Google Play Store version of these apps also had more ratings overall compared to the Apple App Store version.

The applications featured in this study also exhibited many implementation flaws and presented impediments to the user experience. A specific example of this was observed in the “Grounded - Quit Weed” application where the health tab failed to function, displaying a black screen instead. Similarly, the “Marijuana Addiction Calendar” app displayed full-page advertisements frequently. While these could be removed by upgrading to the premium version, free users had a less effective and immersive experience. The “Quit Weed” application, while designed well, also showed advertisements very frequently, which disrupted the user experience.

## DISCUSSION

This study aimed to evaluate the effectiveness of mobile applications for cannabis cessation and assess the current mobile application market in this regard. The results offer valuable insights into the state of the mobile application market for cannabis cessation. The study identified four applications available on both the Apple App Store and Google Play Store for cannabis cessation. Compared to smoking and vaping cessation applications, there were significantly fewer apps for cannabis cessation, and those available had limited use or functionalities.^12,13^

Despite the limited sample size, an association was found between the time of an application’s last update and its MARS score; more recent updates were associated with higher MARS scores. This is likely due to the development of better applications with more features, higher quality experiences, and a focus on target audiences. The MARS analysis suggests a positive relationship between its score and overall application quality, but it is not the sole metric for evaluation. The mobile application market’s lack of standardization leads to a wide variety of features and approaches, making it difficult to draw definitive conclusions about app quality and its relationship to MARS scores. Further research is needed to understand the effectiveness of mobile applications in facilitating cannabis cessation.

Usage tracking was the most commonly employed strategy among both analysed and excluded applications. Most applications primarily tracked the duration of substance use abstinence and other details such as money and time saved. Among the included applications, the “Grounded – Quit Weed” and “Marijuana Addiction Calendar” applications featured a community chat function, while “Marijuana Anonymous” focused on compiling information about in-person community meetings. “Quit Weed” did not have an equivalent. Despite the presence of some features, a greater variety of strategies and tools is needed within the existing set of available applications. Applications should adapt to users’ moods, preferences, and usage to provide personalized healthcare and maximize effectiveness. Recent research has demonstrated the high efficacy of personalized medical applications, such as artificial intelligence chatbots that adapt to users, in changing health behaviours. However, more extensive randomized controlled trials are needed to draw definitive conclusions.^15^

Based on the MARS analysis results, the applications scored the lowest on “information” quality, indicating a lack of evidence-based research within the applications. This is likely because clinicians were not involved in their production, and the focus was on creating engaging and functional experiences rather than information-heavy applications. Nevertheless, recent research has shown that evidence-based mobile applications for behaviours modification, both standalone and as part of an intervention, have advantages over standard health interventions. Despite the overall positive correlation, the heterogeneity of results suggests that more research is needed to determine the specific types of mobile applications and features that yield the most significant benefits.^16^

This study has several strengths, particularly as the first systematic analysis of cannabis cessation applications. It adds to the growing body of research on alcohol, tobacco, and vape cessation applications and informs future research and practice in developing and analysing cessation-focused applications. Based on our findings, we propose several recommendations for improving existing applications and developing future applications to better support the cessation process. Specifically, we suggest that researchers focus on providing adequate, evidence-based information for users and offering personalized experiences. This study lays the groundwork for further research on technology-based interventions, particularly applications and chatbots, to help users quit cannabis. It also contributes to the growing literature on mobile health applications in our healthcare system, particularly concerning substance use and addiction.

It is essential to acknowledge this study’s limitations, especially the small sample size. Evaluating healthcare applications is a recent endeavour, requiring more research and guidance for accurate quality assessment. The rapid pace of application development may have excluded recently added applications from our search and analysis. Moreover, paid features were not evaluated and were beyond the scope of this review. However, free features tend to have the highest usage and accessibility, justifying our approach. Our search was conducted only in Canadian app stores, possibly excluding applications available exclusively in select countries. Furthermore, the applications were only analysed on Apple devices and not on Android devices. This meant some applications may not score comparatively on platforms running alternative operating systems. This was especially concerning as the applications were overall less updated when on the Google Play Store compared to the Apple App Store and upon preliminary review, the applications on the Google Play Store were of significantly lower quality. Lastly, the actual clinical impact of these applications in regard to different users, ethnicities, and cultures was not evaluated. While our analysis did analyse the application’s user interfaces and ease of use through the “Aesthetics” and “Functionality” category, future research should ensure that the applications communicate medical information in an appropriate and easy-to-understand format as well as consider digital and health literacy levels in order to promote increased uptake and reuse rates of the mobile applications.

Overall, the effectiveness of the available mobile applications for cannabis cessation was found to be mixed. The findings suggest that more research is needed to develop evidence-based mobile applications for cannabis cessation to provide individuals with access to high-quality tools to support their cannabis cessation efforts.

## Conclusion

The market for cannabis cessation applications is undersaturated, with the available products having substantial issues that significantly reduce their effectiveness. Given the urgency of these interventions, cannabis cessation applications may prove to be valuable tools in the quitting journey. Therefore, we recommend the development and addition of more robust mobile applications that apply strong, evidence-based approaches toward cannabis cessation.

## Data Availability

All data produced in the present study are available upon reasonable request to the authors

## Notes

### Competing Interest Statement

The authors have declared no competing interest.

### Funding Statement

This study did not receive any funding

## Citations

1. Hawley P, Gobbo M, Afghari N. The impact of legalization of access to recreational Cannabis on Canadian medical users with Cancer. BMC Health Serv Res. 2020 Oct 27;20(1):977. doi: 10.1186/s12913-020-05756-8. PMID: 33109169; PMCID: PMC7590602.

2. Health Canada. Taking stock of progress: Cannabis legalization and regulation in Canada [Internet]. Canada: Government of Canada; 2021 [cited 2023, Apr 30]. Available from: https://www.canada.ca/en/health-canada/programs/engaging-cannabis-legalization-regulation-canada-taking-stock-progress/document.html

3. Health Canada. Canadian Cannabis Survey 2021: Summary [Internet]. Canada: Government of Canada; 2021 [cited 2023, Apr 30]. Available from: https://www.canada.ca/en/health-canada/services/drugs-medication/cannabis/research-data/canadian-cannabis-survey-2021-summary.html

4. Imtiaz S, Nigatu YT, Ali F, et al. Cannabis legalization and cannabis use, daily cannabis use and cannabis-related problems among adults in Ontario, Canada (2001-2019). Drug Alcohol Depend. 2023 Mar 1;244:109765. doi: 10.1016/j.drugalcdep.2023.109765. Epub 2023 Jan 6. PMID: 36652851.

5. Ontario Cannabis Store. A Year in Review 2019-2020. Ontario Cannabis Store; 2019. 15 p.

6. Ontario Cannabis Store. A Year in Review 2020-2021. Ontario Cannabis Store; 2019. 28 p.

7. Canadian Institute for Health Information. Hospital Stays for Harm Caused by Substance Use Among Youth Aged 10 to 24. Ontario: Health Canada; 2019 September. 22 p.

8. Lysyk B. Addictions-Related Treatment Wait Times, Emergency Department Visits and Deaths Rising Despite Increased Spending: Auditor General. Ontario: Office of the Auditor General of Ontario; 2019 December. 2 p.

9. NIDA. Available treatments for marijuana use disorders. [Internet]. National Institute on Drug Abuse (US); [updated 2021 Apr 13; cited 2023 Apr 30]. Available from: https://www.drugabuse.gov/publications/research-reports/marijuana/available-treatments-marijuana-use-disorders.

10. Liu JCJ, Ellis DA. Editorial: Eating in the Age of Smartphones: The Good, the Bad, and the Neutral. Front Psychol. 2021 Dec 6;12:796899. doi: 10.3389/fpsyg.2021.796899. PMID: 34938246; PMCID: PMC8685243.

11. Tofighi B, Chemi C, Ruiz-Valcarcel J, et al. Smartphone Apps Targeting Alcohol and Illicit Substance Use: Systematic Search in in Commercial App Stores and Critical Content Analysis. JMIR Mhealth Uhealth. 2019 Apr 22;7(4):e11831. doi: 10.2196/11831. PMID: 31008713; PMCID: PMC6658280.

12. Thornton L, Quinn C, Birrell L, Guillaumier A, et al. Free smoking cessation mobile apps available in Australia: a quality review and content analysis. Aust N Z J Public Health. 2017 Dec;41(6):625–630. doi: 10.1111/1753-6405.12688. Epub 2017 Jul 27. PMID: 28749591.

13. Sanchez S, Kundu A, Limanto E, et al. Smartphone Apps for Vaping Cessation: Quality Assessment and Content Analysis. JMIR Mhealth Uhealth. 2022 Mar 28;10(3):e31309. doi: 10.2196/31309. PMID: 35343904; PMCID: PMC9002586.

14. Stoyanov SR, Hides L, Kavanagh DJ, et al. Mobile app rating scale: a new tool for assessing the quality of health mobile apps. JMIR Mhealth Uhealth. 2015 Mar 11;3(1):e27. doi: 10.2196/mhealth.3422. PMID: 25760773; PMCID: PMC4376132.

15. Aggarwal A, Tam CC, Wu D, et al. Artificial Intelligence-Based Chatbots for Promoting Health Behavioral Changes: Systematic Review. J Med Internet Res. 2023 Feb 24;25:e40789. doi: 10.2196/40789. PMID: 36826990; PMCID: PMC10007007.

16. Iribarren SJ, Akande TO, Kamp KJ, et al. Effectiveness of Mobile Apps to Promote Health and Manage Disease: Systematic Review and Meta-analysis of Randomized Controlled Trials. JMIR Mhealth Uhealth. 2021 Jan 11;9(1):e21563. doi: 10.2196/21563. PMID: 33427672; PMCID: PMC7834932.

